# Dynamic methods for ongoing assessment of site-level risk in risk-based monitoring of clinical trials: a scoping review

**DOI:** 10.1101/2020.04.01.20049627

**Authors:** William J Cragg, Caroline Hurley, Victoria Yorke-Edwards, Sally P Stenning

## Abstract

**Background/Aims:** It is increasingly recognised that reliance on frequent site visits for monitoring clinical trials is inefficient. Regulators and trialists have in recent years encouraged more risk-based monitoring. Risk assessment should take place before a trial begins in order to define the overarching monitoring strategy. It can also be done on an ongoing basis, in order to target sites for monitoring activity. Various methods have been proposed for such prioritisation, often using terms like ‘central statistical monitoring’, ‘triggered monitoring’ or, as in ICH Good Clinical Practice guidance, ‘targeted on-site monitoring’. We conducted a scoping review to identify such methods, to establish if any published methods were supported by adequate evidence to allow wider implementation, and to point the way to future developments in this field of research.

**Methods:** We used 7 publication databases, 2 sets of methodological conference abstracts and an internet search engine to look for methods for using centrally held trial data to assess site conduct during a trial. We included only reports in English, and excluded reports published before 1996 and reports not directly relevant to our research question. We used reference and citation searches to find additional relevant reports. We extracted data using a pre- defined template. We contacted authors to request additional information about included reports and to check whether reports might be eligible.

**Results:** We included 30 reports in our final dataset, of which 21 were peer-reviewed publications. 20 reports described central statistical monitoring methods (of which 7 focussed on detection of fraud or misconduct) and 9 described triggered monitoring methods. 21 reports included some assessment of their methods’ effectiveness. Most commonly this involved exploring the methods’ characteristics using real trial data with no known integrity issues. Of the 21 with some effectiveness assessment, most presented limited or no information about whether or not concerns identified through central monitoring constituted meaningful problems. Some reports commented on cost savings from reduced on-site monitoring, but none gave detailed costings for the development and maintenance of central monitoring methods themselves.

**Conclusions:** Our review identified various proposed methods, some of which could be combined within the same trial. The apparent emphasis on fraud detection may not be proportionate in all trial settings. Although some methods have self-justifying benefits for data cleaning activity, many have limitations that may currently prevent their routine use for targeting trial monitoring activity. The implementation costs, or uncertainty about these, may also be a barrier. We make recommendations for how the evidence-base supporting these methods could be improved.

## Introduction

Monitoring, a major component of assuring the quality of clinical trials, has traditionally relied on frequent on-site monitoring visits,^1^ particularly to facilitate sometimes extensive source data verification (SDV).^2^ However, it is increasingly recognised that this model may be inefficient and unnecessary in many cases,^3,4^ with trialists questioning the value of 100% SDV.^5–7^ In recent years, regulators^8–10^ and trialists^1,11^ have proposed a risk-based approach to monitoring, whereby monitoring methods, including the frequency and nature of on-site visits, vary across trials depending on the risks specific to each one.

Risk-based monitoring methods can be applied at different stages of a trial. Pre-trial risk assessments can help define the overarching strategies appropriate to the trial’s risks. In some models,^12,13^ this is predominantly a one-off assessment during trial setup. However, it is also possible to modify the monitoring strategy, or build in flexibility and responsiveness, based on changing or emerging risks during the course of the trial.^14^

Risk-based monitoring is often associated with fewer on-site visits than ‘traditional’ monitoring.^15^ Although effective central monitoring methods alone could, in some respects, provide adequate trial monitoring in place of visits, on-site visits offer particular benefits over central monitoring, for example in any activities that require access to site-held source data (such as patients’ medical notes). On-site visits may also be necessary, for example, to investigate potential fraudulent activity. In a risk-based monitoring framework, visits to sites may not be routine, but can be based on assessed risk; we therefore need methods to assess site-level risk on an ongoing basis. We can interpret these methods as assessing the risk of *not going to site now*. If the risk seems too high, a visit – or some other corrective action – is triggered. Methods of this kind have been referred to using various terms, including ‘triggered monitoring’^14^ or, as in ICH GCP guidance, ‘targeted monitoring’,^16^ and may employ data-driven approaches from methods known collectively as ‘central statistical monitoring’,^17^ or more subjective assessments.^14,18,19^

A recent systematic review has established the breadth of tools available to assess overall trial risk (and to use this assessment to define the monitoring strategy) in the setup stage,^20^ but so far there has been no such exercise for methods to assess ongoing site-level risk once a trial has started. We conducted a scoping review^21^ to identify and characterise available methods.

Our aims were 1) for trialists, to establish if any published methods were supported by adequate evidence to support implementation in routine practice and 2) for researchers in this area, to consolidate the existing evidence and point towards future developments in this growing field.

## Methods

We conducted a scoping review to identify methods for using centrally held clinical trial data to assess site-level risk of deviations from Good Clinical Practice (GCP) or the trial protocol, or research misconduct, and thereby to target sites for further monitoring activity. We chose scoping review methodology as we anticipated finding a variety of methods and study types, and we wanted to characterise the extent, range and nature of research activity.^22^ There is no published protocol for this scoping review.

### Eligibility criteria

We defined our eligibility criteria before beginning any searches, with minor refinements (mainly to the exclusion criteria) after having piloted the search strategy.

We included reports:

- Describing methods for using centrally held data (i.e. at the clinical trials unit or other trial coordinating centre) to assess, in ongoing trials, site-level risk of protocol or GCP deviation, risk of data fabrication or research misconduct, or to target sites in some other way for corrective action based on assessed risk (regardless of whether the corrective action involved an on-site monitoring visit or not);
- With methods described in enough detail that we considered them – subjectively – reproducible;
- Either published in peer-reviewed journals or available as grey literature;
- About clinical trials, not limited to trials of Investigational Medicinal Products;
- In English.

We excluded reports:

- Published before 1996 (the year of the first version of ICH GCP Guidance, E6[R1]^23^);
- About quality assurance only in the context of intervention fidelity^24^ or ‘rater differences’^25^ for subjective trial outcome measures;
- About ‘data monitoring’ in general, for example data monitoring committees, unless including methods for assessing site conduct;
- About ‘monitoring’ in any sense other than the GCP sense, e.g. clinical monitoring;
- Focusing only on trial recruitment;
- About more efficient alternatives to standard on-site activity, for example remote source data verification;
- About site selection during trial setup;
- Featuring only opinions, or lacking enough detail allowing theoretical reproduction of methods;
- Reviewing methods presented elsewhere without any new, original methods.

### Information sources and search strategies

#### 1. Database searches

We designed search strategies for the following databases:

- PubMed
- Embase (Ovid)
- Medline (Ovid)
- Web of Science (Clarivate Analytics)
- Cinahl
- Cochrane Central
- Scopus

Full database searches took place on 23 October 2017 (run and extracted by WC). The search strategy for Medline is given in Supplementary Information. We developed our search strategy following review of systematic reviews in this area^1,26^ to identify relevant search terms. We developed the final list through an iterative process in September and October 2017. The final search term combined searches around two concepts: clinical trials (using terms based on those used in a previous systematic review of monitoring methods^26^) and targeted or risk-based clinical trial monitoring. Search terms were modified as required to suit the database being used. No database filters were applied.

Both reviewers (WC, CH) imported results from the seven databases into reference management software and used in-built tools to remove duplicate entries. Both reviewers carried out initial title and abstract screening for relevant reports, producing an initial shortlist of potential papers. These were reduced through review and discussion to a final list of relevant reports for full-text review. We reviewed and discussed these, using full-text reports where possible, to agree a final list of relevant reports. Throughout the process, SS acted as third reviewer where required.

In order to ensure our results were current, this element of the search strategy was repeated on 28 August 2018. WC ran the searches and conducted the title and abstract screening. A shortlist of potentially relevant reports was shared with SS and CH; SS and WC agreed a final list of additional relevant reports from this repeated search.

#### 2. Conference abstracts

We searched for relevant conference abstracts from the first four International Clinical Trials Methodology Conferences (occurring between 2011 and 2017) and all annual meetings of the Society for Clinical Trials since 1996 (initial searches completed on 8 December 2017).

Our methods to search conference abstracts varied depending on the year and the conference: we used website search functions where available and manual PDF searches otherwise. Keywords used for the conference abstract search were based on the key database search strategy terms. These were: ‘monitor’, ‘supervision’, ‘oversight’, ‘risk’, ‘performance’, ‘metric’, ‘quality’, ‘fraud’, ‘fabrication’, and ‘error’.

Both WC and CH performed the abstract searches. This produced an initial shortlist of potentially relevant abstracts. A final list was agreed through discussion, with SS acting as third reviewer where required.

#### 3. Internet searches

We conducted structured searches through Google internet search engine (searches carried out 15-19 December 2017).

Google searches were performed without limitations or use of quotes. Search terms were based on the main database search: ‘Risk based monitoring’, ‘Risk adapted monitoring’, ‘Central monitoring’, ‘Central statistical monitoring’, ‘Triggered monitoring’, ‘Targeted monitoring’, ‘Performance metric’, ‘Site metric’, ‘Key risk indicator’, ‘Site performance’, ‘Centre performance’, ‘Detect fraud’ and ‘Detect fabrication’. We reviewed results on the first 20 pages; if there were no relevant results on any three consecutive pages before this, we stopped reviewing.

WC and CH conducted the searches. Any potential additions to the included list of reports were discussed and agreed, with SS acting as third reviewer where required.

#### 4. References, citations and author contact

To identify other relevant reports, we reviewed references (manually) and citations (using Web of Science) of all papers included or considered for inclusion in the final results, and of review articles relevant to the topic. Whenever required, we contacted report authors to help ascertain if given reports should be included, and to ask about the availability of full-text articles.

### Data collection

We extracted data from full journal articles, where available. We recorded data into an Excel-based tool. WC carried out the final data collection used for this report, with SS double-checking all data for inclusion; consensus was reached on any areas of disagreement. Article authors were contacted (two attempts maximum) for missing descriptive data and further clarifications.

Our data collection template was designed and agreed prior to any data collection, with minor refinement after a first review of all relevant papers (a list of data collection variables is available as Supplementary Information). We collected descriptive data about each of the included reports, including year of publication, type of report and details of the trial(s) it was embedded in. We also looked for any information on cost implications of the proposed methods.

When designing this study, although we predicted we would find a range of methods, we agreed that most of them would in essence address a classification problem, i.e. methods to assign sites a status as ‘concerning’ or ‘not-concerning’, with a ‘true’ deviation status – i.e. confirmed existence of meaningful problems – that could be uncovered by further review. The ‘gold standard’ reference test required to assess true status might be study-specific, but could be on-site monitoring or, if the true status was created through simulation, prior knowledge.

We considered a key measure of the reported methods’ effectiveness to be a demonstrated ability, ideally in a real-life setting, not only to detect ‘true’ sites of concern, but also to show with confidence that sites apparently not of concern are performing well. We therefore aimed to summarise the available information on classification, i.e. any or all of specificity, sensitivity, positive and negative predictive value. We gathered the best reported classification statistics for each method, or, if this was not reported, used available statistics, e.g. number of true and false positives, to calculate these. These calculations were verified by an independent statistician at the Medical Research Council Clinical Trials Unit at University College London.

We did not formally assess the quality of the studies. However, review of the QUADAS-2 tool for quality assessment in diagnostic accuracy studies^27^ informed development of our data collection template.

### Synthesis of results

We did not combine results for individual studies, as it was clear through preliminary review of relevant papers that we would have a variety of study types. Instead, the evidence is summarised descriptively.

## Results

**Figure 1** gives a PRISMA flow diagram^28^ showing the different stages of the review. From the various data sources, we ultimately included 30 reports in our final dataset. 21 of these are peer-reviewed publications. The results are characterised in **Table 1** and listed in full in **Table 2. Figure 2** shows reports by year of publication.

**Table 1:** general characteristics of included studies.

| Characteristic | Number<br>(total=30) | Percentage<br>(%) |
| --- | --- | --- |
| **Publication year** |  |  |
| 1996-2000 | 0 | 0% |
| 2000-2005 | 2 | 7% |
| 2006-2010 | 2 | 7% |
| 2010-2015 | 13 | 43% |
| 2016-2018 | 13 | 43% |
| **Type of source** |  |  |
| Peer-reviewed paper | 21 | 70% |
| Conference abstract or poster | 8 | 27% |
| Thesis | 1 | 3% |
| **Disease setting of trial involved** |  |  |
| Cardiovascular disease | 4 | 13% |
| Emergency medicine | 1 | 3% |
| Haematology | 1 | 3% |
| Infectious diseases | 1 | 3% |
| Mental health | 3 | 10% |
| Neurology | 1 | 3% |
| Oncology | 3 | 10% |
| Ophthalmology | 1 | 3% |
| Renal disease | 1 | 3% |
| Respiratory disease | 1 | 3% |
| Not known or no specific trial involved | 13 | 43% |
| **Geographical setting of trials involved** |  |  |
| Brazil | 1 | 3% |
| International | 7 | 23% |
| Japan | 1 | 3% |
| North America | 4 | 13% |
| UK | 2 | 7% |
| Not known or no specific trial involved | 15 | 50% |
| **Use of Investigational Medicinal Product (IMP) in involved trials** |  |  |
| Involves IMP | 14 | 47% |
| No IMP | 1 | 3% |
| Not known or no specific trial involved | 15 | 50% |
| **Phase of trials involved** |  |  |
| Phase I | 0 | 0% |
| Phase II | 1 | 3% |
| Phase II and III | 1 | 3% |
| Phase III | 9 | 30% |
| Not known or no specific trial involved | 19 | 63% |
| **Status of investigational medicinal product used<sup>a</sup>** |  |  |
| Unlicensed | 0 | 0% |
| Licensed, used outside of its licensed indication | 5 | 17% |
| Licensed, used within its licensed indication | 4 | 13% |
| Not known or no specific trial involved | 22 | 73% |
| **Focus of work<sup>a</sup>** |  |  |
| Central statistical monitoring, focus on fraud or misconduct | 7 | 23% |
| Central statistical monitoring, general | 13 | 43% |
| Triggered monitoring | 9 | 30% |
| Other method(s) for highlighting sites at risk | 2 | 7% |
| **Scope of work** |  |  |
| Description or development of method | 9 | 30% |
| Some assessment of methods' effectiveness | 21 | 70% |

<sup>a</sup> Categories not mutually exclusive

**Table 2:** full listing of all included reports.

| First author, publication year | Type of source | Focus of work | Scope of work |
| --- | --- | --- | --- |
| Agrafiotis, 2018<sup>43</sup> | Peer-reviewed paper | Triggered monitoring | Some assessment of methods' effectiveness |
| Almukhtar, 2015<sup>57</sup> | Conference abstract/poster | Central statistical monitoring, general | Description or development of method |
| Atanu, 2017<sup>58</sup> | Peer-reviewed paper | Central statistical monitoring, general | Description or development of method |
| Bailey, 2017<sup>59</sup> | Conference abstract/poster | Triggered monitoring | Description or development of method |
| Bengtsson, 2017<sup>60</sup> | Thesis | Central statistical monitoring, general | Some assessment of methods' effectiveness |
| Biglan, 2016<sup>33</sup> | Conference abstract/poster | Triggered monitoring | Some assessment of methods' effectiveness |
| Desmet, 2014<sup>39</sup> | Peer-reviewed paper | Central statistical monitoring, general | Some assessment of methods' effectiveness |
| Desmet, 2017<sup>40</sup> | Peer-reviewed paper | Central statistical monitoring, general | Some assessment of methods' effectiveness |
| Diani, 2017<sup>42</sup> | Peer-reviewed paper | Triggered monitoring | Some assessment of methods' effectiveness |
| Djali, 2010<sup>31</sup> | Peer-reviewed paper | Other method(s) for highlighting sites at risk (combines site metric scores directly to flag sites of concern) | Some assessment of methods' effectiveness |
| Dress, 2011<sup>61</sup> | Conference abstract/poster | Triggered monitoring | Description or development of method |
| Edwards, 2014<sup>32</sup> | Peer-reviewed paper | Central statistical monitoring with triggered monitoring | Some assessment of methods' effectiveness |
| Kirkwood, 2013<sup>17</sup> | Peer-reviewed paper | Central statistical monitoring, general | Some assessment of methods' effectiveness |
| Knepper, 2016<sup>19</sup> | Peer-reviewed paper | Central statistical monitoring, focus on fraud or misconduct | Some assessment of methods' effectiveness |
| Knott, 2015<sup>34</sup> | Conference abstract/poster | Central statistical monitoring, general | Some assessment of methods' effectiveness |
| Kodama, 2010<sup>62</sup> | Conference abstract/poster | Central statistical monitoring, focus on fraud or misconduct | Some assessment of methods' effectiveness |
| Lindblad, 2014<sup>35</sup> | Peer-reviewed paper | Central statistical monitoring, general | Some assessment of methods' effectiveness |
| O'Kelly, 2004<sup>36</sup> | Peer-reviewed paper | Central statistical monitoring, focus on fraud or misconduct | Some assessment of methods' effectiveness |
| Pogue, 2013<sup>41</sup> | Peer-reviewed paper | Central statistical monitoring, focus on fraud or misconduct | Some assessment of methods' effectiveness |
| Smith, 2012<sup>30</sup> | Peer-reviewed paper | Other method(s) for highlighting sites at risk (use of "statistical process control methodology" to combine per-site risk indicator scores) | Description or development of method |
| Stenning, 2018<sup>14</sup> | Peer-reviewed paper | Triggered monitoring | Some assessment of methods' effectiveness |
| Taylor, 2002<sup>63</sup> | Peer-reviewed paper | Central statistical monitoring, focus on fraud or misconduct | Some assessment of methods' effectiveness |
| Timmermans, 2016<sup>64</sup> | Peer-reviewed paper | Central statistical monitoring, general | Some assessment of methods' effectiveness |
| Tudur Smith, 2014<sup>18</sup> | Peer-reviewed paper | Triggered monitoring | Description or development of method |
| Valdez-Marquez, 2011<sup>65</sup> | Conference abstract/poster | Central statistical monitoring, general | Description or development of method |
| Valdez-Marquez, 2013<sup>66</sup> | Conference abstract/poster | Central statistical monitoring, general | Description or development of method |
| van den Bor, 2017<sup>52</sup> | Peer-reviewed paper | Central statistical monitoring, focus on fraud or misconduct | Some assessment of methods' effectiveness |
| Whitham, 2018<sup>67</sup> | Peer-reviewed paper | Triggered monitoring | Description or development of method |
| Wu, 2011<sup>38</sup> | Peer-reviewed paper | Central statistical monitoring, focus on fraud or misconduct | Some assessment of methods' effectiveness |
| Zink, 2018<sup>68</sup> | Peer-reviewed paper | Central statistical monitoring, general | Some assessment of methods' effectiveness |

**Figure 1.**
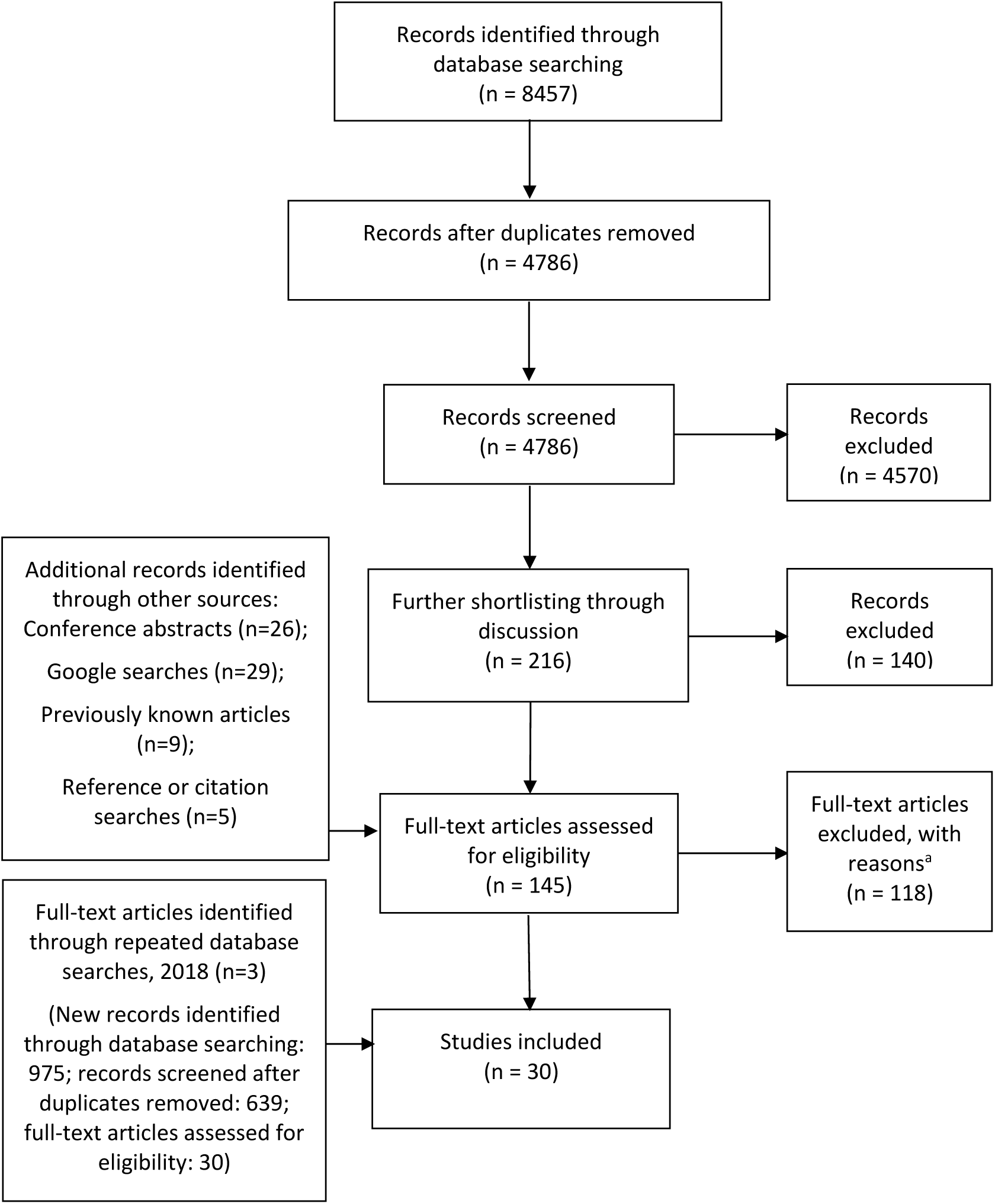
PRISMA flow diagram. ^a^ Reasons: no relevant methods presented (n=28); no novel methods presented (e.g. review article; n=28); method to measure variation between trial sites but no ‘flagging’ of sites of concern (n=25); abstract only and not enough detail to confirm relevance (n=10); duplicate or abstract where full paper also available (n=8); grey literature not considered to present reproducible methods (n=5); not about ‘monitoring’ according to ICH Good Clinical Practice definition (n=5); trial-level assessment only, not site-level (n=4); focus on consistency of outcome assessment only (n=4); method from observational study only, not clinical trial (n=1).

**Figure 2.**
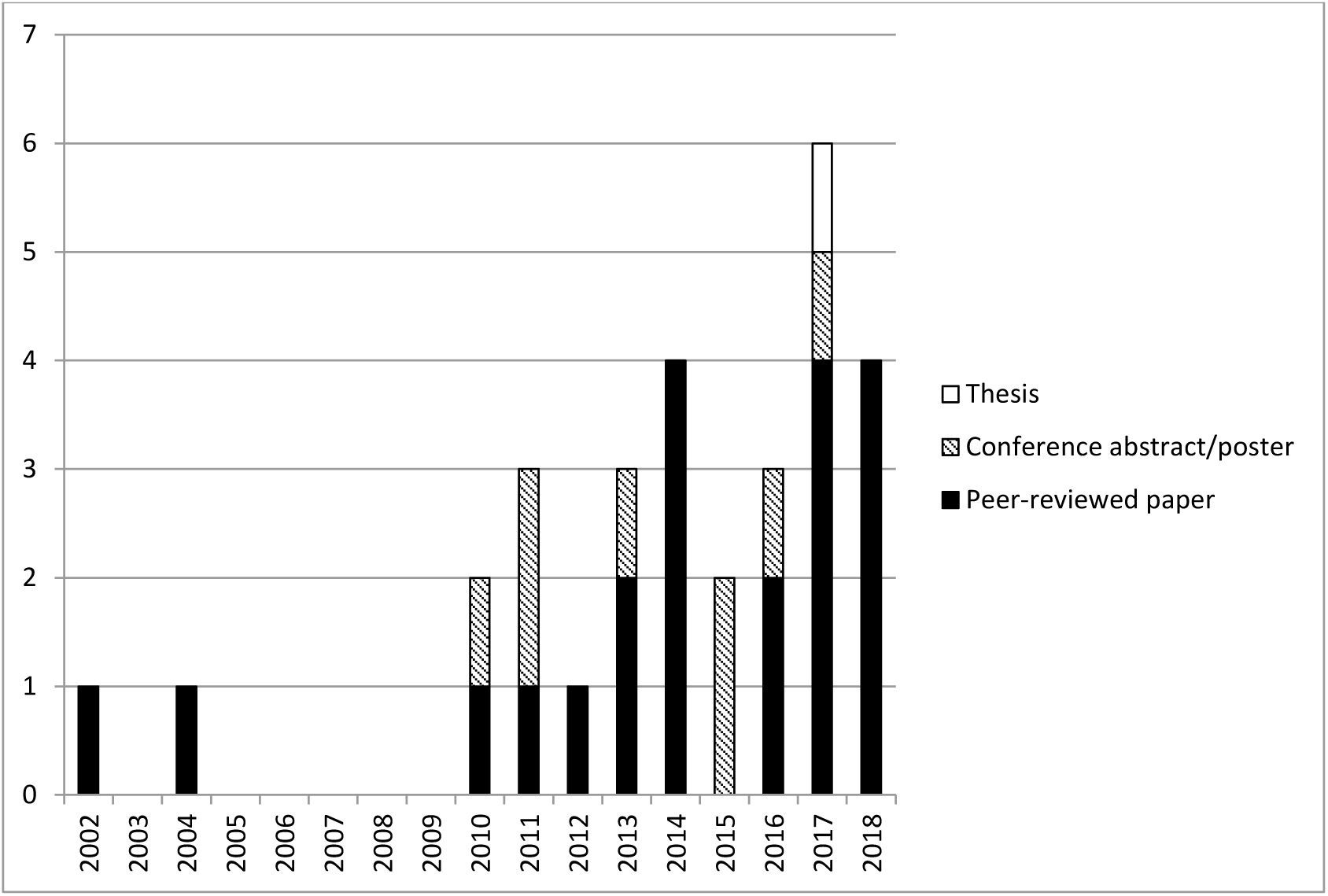
Publications by year and type.

When specific trials were mentioned, they involved various health conditions and were in various geographical settings. Information on trial intervention was not often available, but where it was, methods had most often been used in phase III trials of investigational medicinal products (IMP). The IMP risk category,^29^ when known, was either ‘licensed and used within its licensed indication’, or ‘licensed and used outside its licensed indication’ (i.e. we found no reports involving trials of unlicensed IMPs).

We classified 20/30 of our results as central statistical monitoring methods, of which 7 focussed on detection of investigator fraud or research misconduct. We classified 9, including one of the 20 that used central statistical monitoring, as ‘triggered monitoring’, i.e. review of each trial site against pre-set thresholds in key performance metrics, usually without any statistical testing. A final 2 we could not fit into either of these categories; these involved using measured site metrics to directly compare sites against one another.^30,31^

21/30 reports included some assessment of the effectiveness of the methods; these are summarised in **Table 3**. The most common experimental designs were to explore the methods’ characteristics using real trial data with no known integrity issues (n=9), and simulating data integrity problems at sites within real trial datasets then using the method to try to identify the problem sites (n=6).

**Table 3:**
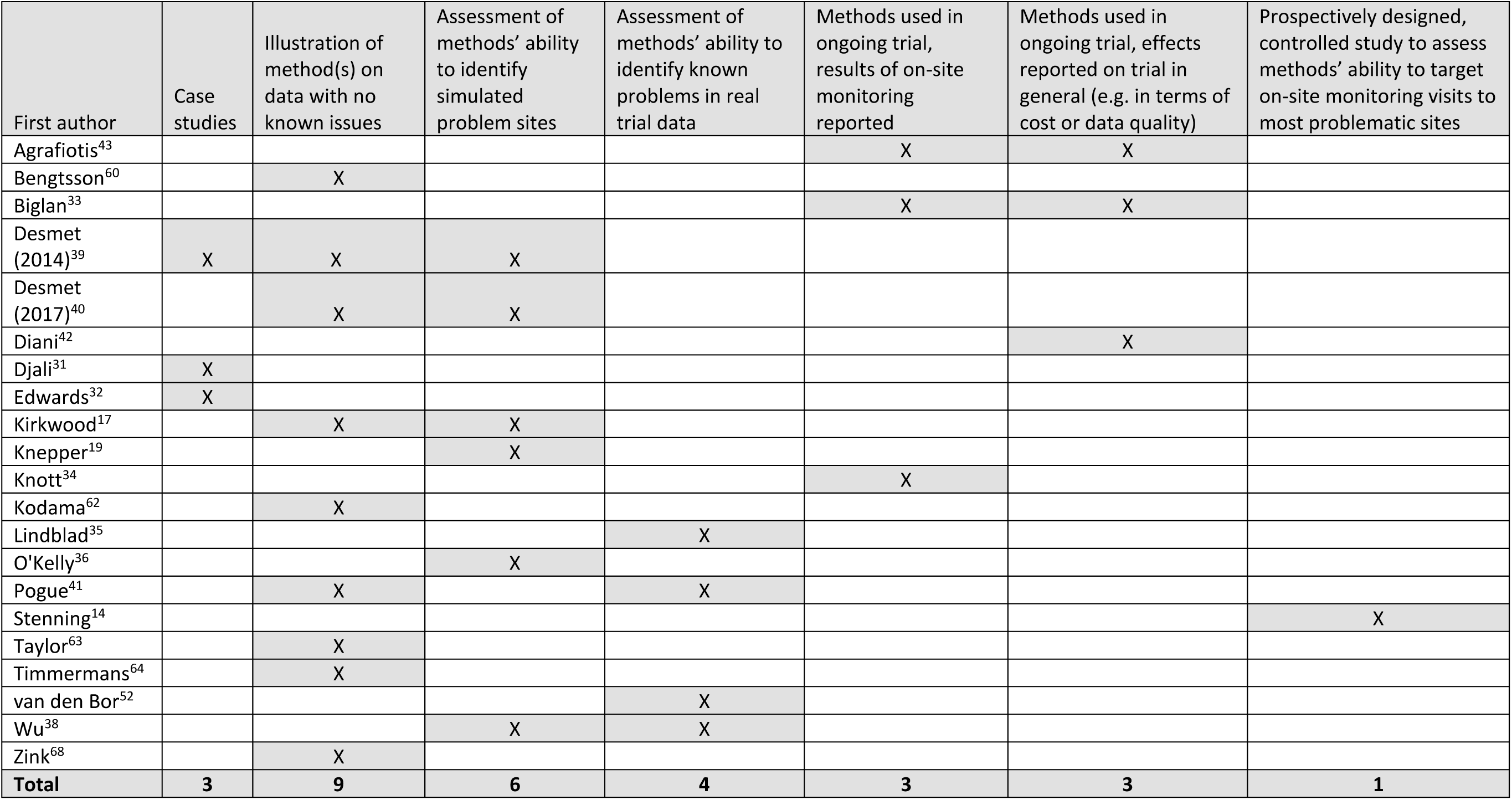
Types of assessments and evidence presented by reports that included some assessments of their methods’ effectiveness.

Of the 21 reports, 9 had no information about sites’ ‘true’ status, i.e. whether the problems identified through central monitoring constitute meaningful problems (either recorded through on-site monitoring or audit activity, or known because statuses were created through simulation). One report^32^ only contained case studies, i.e. partial and selective reporting. Seven^14,33–38^ had partial information, e.g. some of sites’ true statuses were reported, but not all. Two explored classification ability through extensive simulation,^39,40^ and 2 had detailed information from a limited set of scenarios on the number of true and false positives and negatives.^19,41^ The best reported or deducible classification ability for the 11 papers with at least some information on sites’ ‘true’ status is shown in **Table 4;** in many cases this is limited by considerable amounts of missing or unclear data.

**Table 4:** best reported information on methods’ classification ability, where available or deducible.

| First author | Available information on methods' classification abilities | Definition of 'positive' centres | 'True' test status: real or simulated? | Test for 'true' centre status | Sensitivity<sup>a</sup> | Specificity<sup>b</sup> | Positive Predictive Value<sup>c</sup> | Negative Predictive Value<sup>d</sup> |
| --- | --- | --- | --- | --- | --- | --- | --- | --- |
| Biglan<sup>33</sup> | Partial ('true' status known for only one centre; total number of centres not known) | Not clearly defined<sup>e</sup> | Real | On-site monitoring | Unavailable due to limited data; report states that one 'low-risk' centre was visited and considered to be misclassified (i.e. should have been 'medium risk' or 'high risk'). However, total number of sites classified and visited (overall and within each risk category) is not known. |  |  |  |
| Desmet (2014)<sup>39</sup> | Explored through simulation | Presence of atypical data | Simulated | Known because simulated | Dependent on simulation scenario; no specific figure given |  |  |  |
|  | Detailed information (vital signs data used as illustrative example) | Presence of atypical data | Real | Unclear ('closer inspection') | **Reported:** 83% (10/(10+2)) | **Reported:** 99% (204/(204+2)) | **Calculated:** 83% (10/(10+2)) | **Calculated:** 99% (204/(204+2)) |
| Desmet (2017)<sup>40</sup> | Explored through simulation | Presence of atypical data | Simulated | Known because simulated | **Reported:** dependent on simulation scenario; no specific figure given | **Reported:** median specificity varied from 98%-100% depending on scenario | Not reported and not possible to calculate (results of many simulations presented) |  |
| Knepper<sup>19</sup> | Detailed information | Presence of fabricated data | Simulated with physician input | Known because simulated | **Reported:** best result from 4 scenarios (study 1): 86% (6/(6+1)) | **Reported:** best result from 4 scenarios (study 1a): 87% (148/(148+23))<sup>f</sup> | **Reported:** best result from 4 scenarios (study 2a): 27% (3/3+8) | **Reported:** best result from 4 scenarios (study 1): 99% (132/132+1) |
| Knott<sup>34</sup> | Partial (total number of sites not reported but likely more than number whose results reported; 'true' status of any unreported centres not known) | Presence of any findings | Real | On-site monitoring | **Calculated:** 85% (11/(11+2)) | **Calculated:** 88% (7/(7+1)) | **Calculated:** 92% (11/(11+1)) | **Calculated:** 78% (7/(7+2)) |
|  |  | Presence of findings 'indicative of sloppy practice' (clearer definition not reported) | Real | On-site monitoring | **Calculated:** 83% (10/(10+2)) | **Calculated:** 78% (7/(7+2)) | **Calculated:** 83% (10/(10+2)) | **Calculated:** 78% (7/(7+2)) |
|  |  | Presence of serious findings (clearer definition not reported) | Real | On-site monitoring | **Calculated:** 100% (1/1+0) | **Calculated:** 45% (9/(9+11)) | **Calculated:** 8% (1/(1+11)) | **Calculated:** 100% (9/(9+0)) |
| Lindblad<sup>35</sup> | Partial ('true' status known only at 21/413 centres) | Presence of serious problems | Real | Regulatory inspection | **Reported:** 83% (5/((5+1)) | Cannot be calculated without making assumptions about the 392/413 sites with unknown 'true' status |  |  |
|  |  | Presence of minor problems | Real | Regulatory inspection | **Reported:** 89% (8/(8+1)) |  |  |  |
|  |  | Presence of any problems | Real | Regulatory inspection | **Reported:** 87% (13/(13+2)) |  |  |  |
| O'Kelly<sup>36</sup> | Detailed information, but sample of data from trial <sup>g</sup> | Presence of fabricated data | Simulated with physician input | Known because simulated | **Calculated:** 33% (1/(1+2)) | **Calculated:** 95% (18/(18+1)) | **Calculated:** 50% (1/(1+1)) | **Calculated:** 90% (18/(18+2)) |
| Pogue<sup>41</sup> | POISE trial data: detailed information from all sites with >= 20 randomisations | Presence of fabricated data | Real | On-site monitoring | **Reported:** different models and different thresholds give different pros and cons in terms of classification. Models 1, 3 and 5 all have at least some scenarios where both specificity and sensitivity > 80%. (Model 1 and 5, risk score >=7; Model 3, risk score >=5) |  |  |  |
|  | HOPE trial data: summary information from all sites with >= 20 randomisations | Presence of fabricated data | Real | On-site monitoring | N/a (no true positives) | **Reported:** model 1: 99% (178/(178+2)) | **Calculated:** all models: 0% (no true positives, so any positives are false) | **Calculated:** all models: 100% (no true positives, so all negatives are true negatives) |
| Stenning<sup>14</sup> | Partial (only sample of negative-testing sites visited, although the study design aimed to control for this) | Presence of ≥ 1 serious (Major or Critical) finding | Real | On-site monitoring | **Calculated:** primary analysis: 52% (37/(37+34)) | **Calculated:** primary analysis: 62% (8/(8+5)) | **Reported:** primary analysis: 88% (37/(37+5)) | **Calculated:** primary analysis: 19% (8/(8+34)) |
|  |  |  |  |  | **Calculated:** secondary analysis excluding re-consent findings: 59% (36/(36+25)) | **Calculated:** secondary analysis excluding re-consent findings: 74% (17/(17+6)) | **Reported:** secondary analysis excluding re-consent findings: 86% (36/(36+6)) | **Calculated:** secondary analysis excluding re-consent findings: 40% (17/(17+25)) |
|  |  |  |  |  | **Calculated:**<br>secondary analysis excluding all consent findings: 60% (29/(29+19)) | **Calculated:**<br>secondary analysis excluding all consent findings: 64% (23/(23+13)) | **Reported:**<br>secondary analysis excluding all consent findings: 69% (29/(29+13)) | **Calculated:**<br>secondary analysis excluding all consent findings: 55% (23/(23+19)) |
| van den Bor<sup>37</sup> | Partial in paper, but authors confirmed that trial implemented source data verification for all sites (personal communication) | Presence of fabricated data | Real | On-site monitoring | Various situations presented, with different implications for classification ability.<br>Median false positives below 10% for all scenarios, lower with higher m-constant; in various situations (combinations of specific m-constants with specific scenarios), the fraudulent centre is flagged $\geq 3$ times (authors' proposed threshold) 100% of the time.<br>Some scenarios have 100% highlighting of fraudulent centre and very low false positive rate - e.g. scenario 1, m=20, scenario 2, m=2-, scenario 3, m=20 (all with false positive rate of 2%) |  |  |  |
| Wu<sup>38</sup> | Partial (15/17 sites have unknown 'true' status) | Presence of fabricated data | Real | Auditing | Results presented narratively via a number of scenarios.<br>For 'angular clustering', fourth scenario (correlation 0.7, 3 outliers) results in sensitivity, specificity, positive and negative predictive values all $\geq 98\%$. For 'neighborhood clustering', specificity in all scenarios is $\geq 94\%$ and second scenario (variances 0.45, 3 outliers, cluster size 27) results in sensitivity, specificity, positive and negative predictive values all $\geq 50\%$. |  |  |  |

<sup>a</sup> Number of correctly flagged problem sites / (number of correctly flagged problem sites + sites incorrectly *not* flagged as concerning); thick border used to highlight results more than or equal to 90%.
<sup>b</sup> Number of sites correctly flagged as not concerning / (number of sites correctly flagged as not concerning + sites incorrectly flagged as concerning); thick border used to highlight results more than or equal to 90%.
<sup>c</sup> Number of correctly flagged problem sites / (number of correctly flagged problem sites + sites incorrectly flagged as concerning); thick border used to highlight results more than or equal to 90%.
<sup>d</sup> Number of sites correctly flagged as not concerning / (number of sites correctly flagged as not concerning + sites incorrectly *not* flagged as concerning); thick border used to highlight results more than or equal to 90%.
<sup>e</sup> One 'positive' centre is described as "reveal[ing that] RBM was not assessing risk sufficiently to drive monitoring decisions"
<sup>f</sup> Publication incorrectly rounds this to 86%.
<sup>g</sup> Approximately one third of sites included from a trial; also some uncertainty about total number of sites (sometimes reported as 21, sometimes 22; used 22 for calculations given here as this is figure in Results section)

Some papers report on actual or theoretical cost savings from reduced on-site monitoring,^33,42,43^ and others comment on the risk of incurring costs if their proposed central monitoring method identifies sites that do not in fact have meaningful problems (i.e. false positives).^19,37^ However, no papers give detailed costings for the development, implementation and maintenance of the central monitoring methods themselves.

## Discussion

We conducted a scoping review to identify published methods for assessing the risk of not taking corrective action at trial sites at a given time. Although our search looked for reports from any time after 1995, over half of our results are from after 2013, highlighting the recent growth of risk-based monitoring concepts. Host trials for these methods were most often phase III trials of licensed investigational medicinal products, i.e. somewhat lower risk compared to earlier phase trials of newer treatments.^29^ Around a third of our results were not full, peer-reviewed reports, reflecting a wider problem with evidence supporting trial conduct methods, i.e. that researchers can feasibly produce posters and abstracts for conferences, but may not have time or incentive to produce comprehensive reports.^44^

Identified methods were mainly in two broad categories (with some overlap). Most were about central statistical monitoring, which uses statistical testing of all or a subset of trial data items to compare sites and identify atypical trial centres. A minority described triggered monitoring techniques, whereby sites are assessed against pre-specified site metric threshold rules (usually binary), with sites meeting the greatest number of ‘triggers’ being considered the most concerning. Several authors note that central statistical monitoring needs sufficient overall and per-site sample sizes for adequate statistical power.^17,19,37^ Triggered monitoring, however, can be used at any stage of a trial’s recruitment (especially with trigger rules based on single instances of a given protocol violation, for example). We therefore suggest use of the techniques is not mutually exclusive.

Although the reports on central statistical monitoring described a range of methods, there were some commonalities. Previous papers have reviewed these methods in more detail.^17,41,45–47^

Nearly half of the central statistical monitoring reports focused on identification of data fabrication. The possibility of fraud is a serious concern to trialists and a threat to wider trust in science.^48^ It was possibly an important factor in establishing 100% on-site verification of trial data as a common monitoring approach.^49,50^ This may help explain the prevalence of reports about fraud detection, as some may see the priority in risk-based monitoring to be establishing its fraud detection ability compared with 100% source data verification. However, although the incidence of data fraud is difficult to quantify, cases of extensive data fabrication appear rare enough to have individual notoriety.^51^ Further, methods to detect fraud are necessarily rather selective, and therefore may not alone be suitable for trialists looking to detect more common, lower-level data integrity issues.

We collected data on how the proposed methods we identified had been evaluated. A number of reports only presented proposed, untested methods, or only selected case studies to demonstrate the methods’ performance. Of those that presented more detailed evaluation, a common limitation was that the ‘true’ status both of identified problem sites and sites apparently not of concern was often not available, or only partially available. It was therefore difficult to know if the ‘concern’ status of sites in central monitoring results represented meaningful problems or not. In addition, a number of studies use simulation to create ‘true’ sites of concern; these raise the additional question of whether these simulations reflect real-life issues, though the involvement of clinicians (i.e. those who would provide real-life trial data) in the simulation process of some reports^19,36^ is reassuring.

Clearly, a balance must be struck between minimising cost and adequately mitigating key risks. Many false positives could be expensive (e.g. additional on-site monitoring visits for no gain), but many false negatives could be catastrophic (e.g. missing serious risks to trial participants’ safety). Some authors do acknowledge this, and even suggest that a high false positive rate could be acceptable in a method used to broadly target additional scrutiny at particular sites.^38^

It is important to recognise the limitations of the available ‘gold-standards’ in the classification of sites. When methods are tested using simulated or real-but-adjusted data, it may be difficult to know how well these accurately recreate real-life situations. When central monitoring methods are tested in real, ongoing trials, on-site monitoring may be an imperfect reference test, in that it may not be able to identify all problems. By contrast, it is clear that central monitoring, with its enhanced inter- and intra-site review, can identify issues that a single team at one site for a limited time might not.^47^

It could be argued that at least some of the methods we have identified do not need extensive evaluation because they prove their own worth. For instance, they help identify outliers that in some cases are self-evidently meaningful problems to resolve. We acknowledge that some central monitoring activities identify ‘known’ problems (e.g. identifying weekend visit dates, which in most cases are unlikely to be correct) and are valuable for data cleaning purposes. However, we were specifically interested in the more nuanced use of these methods to identify sites of ‘concern’, at which monitoring activity may be targeted, and consequently sites ‘not of concern’, monitoring of which may be reduced or omitted. This element does need adequate proof before wider adoption and, if it is shown to be effective, could have significant benefits. In light of the limitations we have described here, we do not believe any methods have yet fully demonstrated that they should be adopted more widely as a means to limit on-site monitoring of sites deemed, based on review of centrally-held data, not to be of concern.

Aside from some comments on the potential cost of investigating false positive central monitoring results,^19,52^ the reports we identified contained limited information on the cost of developing and implementing their methods. As well as uncertainty about how to develop relevant methods, uncertainty or concern about costs involved is a substantial barrier to adoption of risk-based monitoring.^53^

Through our various searches, we identified 24 commercial companies which, we were at least reasonably certain, had a method relevant to our search (data not shown). Only a small proportion of these had published details of their methods in peer-reviewed journals, although just over half had some detail in grey literature sources. This highlights another difficulty in disseminating new trial conduct methods, i.e. that they may be commercially sensitive and, unlike evidence about treatments being tested in trials, there is no compulsion to publish new evidence. It is somewhat contradictory that while risk-based monitoring has come about partly to reduce costs of trials, some risk-based monitoring methods are available only for a fee.

Further work is needed to fully demonstrate the effectiveness of these dynamic site risk assessment methods which, alongside pre-trial risk assessments, form the core of risk-based monitoring. We therefore recommend the following:

1. **Coordinate research efforts**. From the scoping review and contact with report authors, it was clear that various small research projects relevant to this topic were ongoing, but mostly in isolation. Researchers in this area should take stock of existing research, and set clear priorities to ensure research time is well-spent.
2. **Standardise monitoring studies**. Core outcome sets^54^ or other mechanisms to standardise studies about monitoring would improve study quality and may facilitate cross-study evidence synthesis.
3. **Share evidence**. Time, commercial sensitivity and perceived reputational risk could all be barriers to publishing evidence about monitoring practices. However, additional, publicly-available evidence to support best monitoring practice will allow trialists in all settings to adopt new methods with confidence.
4. **Publish full papers**. Conference abstracts and posters are a useful way to disseminate basic information about new ideas, but rarely have enough detail to allow replication or robustly demonstrate effectiveness. As this emerging field cannot be built on abstracts alone, we encourage researchers to publish full, peer-reviewed papers about their monitoring methods.
5. **Look to combine complementary methods**. Although work has been done on a number of distinct risk-based monitoring methods, an optimal monitoring plan might involve a combination of these, including both central statistical monitoring and triggered monitoring. A collaborative approach to combining existing methods could help develop and test such an idea.

We acknowledge several limitations. Our database searches identified relevant material from disparate locations, including abstracts in conferences in unrelated research fields. It is possible that other abstract collections include relevant material, but it was not feasible to find and hand-search all of these. Although the internet searches made little contribution to the final list of included reports, they may have been limited by known reproducibility problems.^55^

Scoping review methodology advises that relevant experts in a field are surveyed to help identify other relevant work.^56^ We have not formally done this. We have, however, contacted most authors of included reports for clarifications, and this has not highlighted any additional relevant reports.

Although we pre-specified our aims and our eligibility criteria, we added some detail to our exclusion criteria during the course of our review to help us decide about certain reports. It is possible that other researchers repeating the same review might result in a slightly different list, but we believe this might only affect the ‘method-only’ papers, which do not form the key part of our conclusions. The comprehensive nature of our search strategy, including review of reference and citation lists, gives us confidence that our report is a sound overview of the state of the evidence in this research area.

We have not performed a formal quality assessment of reports we found, however, this is considered by some to be unnecessary in scoping review methodology.^22^ There is also no validated way to review the quality of risk-based monitoring studies, although we used the QUADAS-2 tool, designed to assess the quality of diagnostic studies, to inform our data collection template.

Finally, we acknowledge that some time has now passed since we first conducted our search for relevant evidence. Conscious of this, we repeated the main database search in 2018 (albeit with only one author conducting title and abstract screening) and added three relevant reports. We are not aware of any research published since then that might change our overall conclusions. If evidence is now available that addresses the limitations we have highlighted in the existing literature, we would certainly consider this a positive development.

Our scoping review highlighted some promising evidence for risk-based monitoring in ongoing trials. However, currently published methods may not yet have demonstrated their efficacy or cost-effectiveness well enough for trialists to implement them with confidence as a means to target or omit on-site visits. A more coordinated, collaborative and transparent approach to developing and sharing evidence in this field, including industry and academic partners, could help it grow beyond its current nascent state, and could contribute to risk- based monitoring more quickly entering routine practice.

## Data Availability

The data supporting this work are available on reasonable request. Please contact the corresponding author in the first instance.

## Acknowledgements

We would like to thank the authors of reports – some referenced in this manuscript and some ultimately not – for their time in answering our questions and discussing their work with us. We would also like to thank: systematic reviewers at Medical Research Council Clinical Trials Unit at University College London (MRC CTU at UCL) for their advice; Meredith Martyn for verifying the classification statistics, Sharon B Love for project support and Matthew R Sydes for advice, project support and review of this manuscript.

## Supplementary information

1. Completed PRISMA-ScR Checklist
2. Search strategy from Medline
3. Annotated list of data collection variables

